# The effect of sample medication use on subsequent anti-VEGF agent selection for neovascular age-related macular degeneration

**DOI:** 10.1101/2021.07.27.21261104

**Authors:** Karen M. Wai, Tedi Begaj, Sachi Patil, Evan Chen, John B. Miller, Jan Kylstra, Mary E. Aronow, Lucy H. Young, Rachel Huckfeldt, Deeba Husain, Leo A. Kim, Demetrios G. Vavvas, Dean Eliott, Shizuo Mukai, Evangelos S. Gragoudas, Nimesh A. Patel, Lucia Sobrin, Joan W. Miller, Ravi Parikh, David M. Wu

## Abstract

**Purpose:** To examine the effect of medication sample use (ranibizumab or aflibercept) on future anti-vascular endothelial growth factor (VEGF) agent selection in neovascular age-related macular degeneration (nvAMD).

**Design:** Retrospective cohort study.

**Methods:** nvAMD patients who underwent an initial anti-VEGF injection with a sample medication were compared to nvAMD control patients who never received a medication sample. Charts from 2017 through 2020 were reviewed for data regarding demographics, anti-VEGF agent selection, and visual acuity outcomes for both groups. Anti-VEGF agent selection for the first four injections and at one year were examined in both the sample and control groups.

**Results:** Adherence to the initial agent was high between first and subsequent injections (2^nd^, 3^rd^, 4^th^ injection, and 1 year) in both sample (96.2%, 95.9%, 91.9%, 93.4%, respectively) and control groups (98.1%, 94.2%, 94.9%, 87.8%, respectively). Bevacizumab usage was significantly lower among eyes receiving samples relative to controls at the second (1.9% vs. 38.7%, p<0.001), third (3.1% vs. 41.3%, p<0.001), fourth injections (4.7% vs. 40.4%, p<0.001), and at 1 year (0% vs. 33.8%, p<0.001). Aflibercept usage was significantly higher in sample eyes relative to controls at the second (78.3% vs. 43.4%, p<0.001), third (76.3% vs. 41.5%, p<0.001), and fourth injections (76.7% vs. 43.4%, p<0.001), and at 1 year (77.0% vs. 52.7%, p<0.001).

**Conclusions:** Eyes receiving a sample anti-VEGF agent (ranibizumab or aflibercept) for their initial injection were less likely to receive bevacizumab at future visits relative to eyes that did not receive an anti-VEGF sample, even after one year of treatment.

## Introduction

Neovascular age-related macular degeneration (nvAMD) is a rapidly progressive disease that requires early initiation of treatment to prevent irreversible anatomical and functional loss. A recent study demonstrated that a 2-week delay in the administration of anti-vascular endothelial growth factor (anti-VEGF) after initial diagnosis is associated with worse visual outcomes.^1^ Additional research has demonstrated improved visual outcomes for nvAMD with a shorter time span between symptom presentation and treatment.^2^ Unfortunately, the initiation of treatment for patients with nvAMD is often delayed due to insurance prior-authorization (PA) requirements. PAs can inconvenience patients, complicate the timely initiation of care and have been shown to increase rates of treatment abandonment in other specialties.^3,4^

Medication samples represent a convenient, no-cost option to immediately provide treatment to patients with nvAMD while PA requests are pending. However, it is well established in medicine that samples can influence long-term physician prescribing behaviors.^5^ Additionally, sample availability often skews towards expensive branded medications.^5,6^ For nvAMD treatment, Bevacizumab (Avastin; Genentech, South San Francisco, CA) is the least expensive anti-VEGF agent. Initially a chemotherapy medication, it was repurposed as an intravitreal injection and used as an off-label drug by ophthalmologists, which enables a relatively low-cost of the drug at approximately $50 per 1.25 mg dose when mixed by a local compounding pharmacy.^7^ Because of the nature by which it is supplied, there are no samples of bevacizumab. Brand name on-label medications including ranibizumab (Lucentis; Genentech, South San Francisco, CA) and aflibercept (Eylea; Regeneron, Tarrytown, NY) have higher costs, with a single vial costing roughly $1716.55 (0.5mg/0.05ml) and $1876.88 (2.0mg/0.5ml) respectively.^8^ Both ranibizumab and aflibercept are approved by the Food and Drug Administration (FDA) for the treatment of nvAMD, and available as samples as a courtesy from their respective pharmaceutical companies.

Increasing drug costs are a significant concern as utilization of anti-VEGF agents is rising in the United States. Furthermore, nvAMD accounts for nearly two thirds of anti-VEGF utilization.^9^ With the large price discrepancies between anti-VEGF medication options and the chronic nature of anti-VEGF treatment, the availability of samples may have significant ramifications on administration patterns and cost of care. Anti-VEGF medications are administered by the physician in clinic (i.e. Medicare Part B not D) under a model where the initial capital cost is placed on the physicians/clinic in the “buy and bill” model.^6,10^ Thus, when presented with a new nvAMD patient, the physician must either defer treatment until prior authorization can be obtained, treat with a medicine (such as bevacizumab) and shoulder the cost should reimbursement later be denied, or if available, use an on-label sample which allows prompt treatment at no initial cost. There is no pre-existing literature on intravitreal anti-VEGF sample usage among ophthalmologists and how it may relate to future anti-VEGF selection. Therefore, the goal of our study is to determine how initial utilization of anti-VEGF samples affects subsequent anti-VEGF agent selection for patients with nvAMD.

## Methods

A retrospective cohort study was conducted at a single institution, Massachusetts Eye and Ear (MEE). This study was approved by the institutional review board of the Massachusetts General Hospital and Partners Healthcare (IRB # 2021P000301) and is compliant with the Declaration of Helsinki and Health Insurance Portability and Accountability Act regulations.

Patients with an ICD-9 or ICD-10 code for nvAMD who underwent their first anti-VEGF injection at MEE between January 2017 and December 2020 were identified. Eyes were included if they received a minimum of 2 anti-VEGF injections. Only one eye from each patient was included in the study; if both eyes met inclusion criteria, one eye was selected randomly to be included.

Billing codes were used to identify if a patient received a sample anti-VEGF medication with ranibizumab or aflibercept as their first injection at MEE. Patients were excluded if there was any history of prior injections at an outside institution or if there were any pre-existing indications that may have required anti-VEGF therapy (e.g. diabetic macular edema, retinal vein occlusion with macular edema). One-hundred and six patients fit this inclusion criteria for the sample group.

The control group was comprised of patients with nvAMD who did not receive a sample medication for their first injection, but rather an insurance-approved anti-VEGF agent. To account for variation between the sample and control patients among different MEE attendings at three MEE satellite facilities where samples are available, control patients were matched by both treating attending and location of injection to the sample population. Only patients from MEE attendings who injected both samples and insurance-covered medications, and who injected both on and off-label medications were included in the study. The control population consisted of several hundred patients; thus, a random sequence generator was used to randomize the patients, and the first 106 patients who met inclusion criteria were included in the study. Sample patients had similar exclusion criteria applied to them. Patients seen at MEE satellite locations who did not have access to sample injections were also excluded from the study.

For all patients who met inclusion criteria, the following was collected: patient demographics, date of diagnosis relative to first injection, anti-VEGF agent selected for their first four injection visits and whether the agent was a sample or insurance-covered, visual acuities at each visit injection visit, and what agent was used at the one year time point if available. Date of first retina specialist evaluation for nvAMD was recorded. Insurance data was collected and divided into Medicare vs. Private/Medicare Advantage; this stratification was performed as Medicare does not typically require prior authorization for anti-VEGF injections. Adherence at each visit was calculated as a percentage of patients who received the same agent at that particular injection visit that they received at their first injection visit.

For statistical analysis, multivariate analysis taking into account gender, age, race, insurance, and location was used to compare the baseline demographics between the two groups. The proportion of patients receiving each agent was calculated at each of the time points: initial injection, second injection, third injection, fourth injection, and one year. Chi-square test for independence of proportions was used to compare anti-VEGF agent selection at each time point between the groups. An alpha of 0.05 was used as the threshold for statistical significance. Independent t-tests were used to compare time from visit with a retina specialist to first injection. Snellen acuities were converted to LogMAR, and acuities were compared between groups with independent t-tests. P values less than <0.05 were considered significant.

## Results

### Baseline demographics

One hundred and six eyes from 106 patients were identified who received a medication sample (either ranibizumab or aflibercept) as their initial anti-VEGF injection. Of these 106 sample patients, 68 (64.2%) were female. The average age was 79.7 years [standard deviation (SD): 8.2 years], and 87.7% of patients were white. The control population consisted of patients who received only insurance-covered anti-VEGF injections for nvAMD, without any sample utilization in their treatment course. Of these 106 control patients, 69 (65.1%) were female with an average age of 79.2 years (SD: 8.5 years). The majority of patients (82.1%) were white (**Table 1**).

**Table 1:** Baseline demographics of sample and control group.

| **Table 1: Baseline demographics of sample and control group** |  |  |  |
| --- | --- | --- | --- |
|  | **Sample**<br>**population**<br>**(n=106)** | **Control**<br>**population**<br>**(n=106)** | **P value** |
| **Gender** |  |  |  |
| Male | 38 (35.8%) | 37 (34.9%) | 0.53 |
| Female | 68 (64.2%) | 69 (65.1%) |  |
| **Age, mean (SD)** | 79.7 (8.2) | 79.2 (8.5) | 0.89 |
| **Race** |  |  |  |
| White | 93 (87.7%) | 87 (82.1%) | 0.10 |
| Black | 4 (3.8%) | 0 (0%) |  |
| Asian | 0 (0%) | 4 (3.8%) |  |
| Hispanic | 0 (0%) | 0 (0%) |  |
| Unavailable | 9 (8.5%) | 15 (14.1%) |  |
| **Insurance** |  |  |  |
| Medicare | 42 (39.6%) | 78 (73.6%) | 0.06 |
| Medicare Advantage/Private | 64 (60.4%) | 28 (26.4%) |  |
| **Location of first injection** |  |  |  |
| Site 1 | 29 (27.4%) | 29 (27.4%) | 0.93 |
| Site 2 | 18 (17.0%) | 18 (17.0%) |  |
| Site 3 | 59 (55.7%) | 59 (55.7%) |  |

With multivariate analysis, there were no significant baseline differences between the sample population and control population in gender (p=0.53), age (p=0.89), race (p=0.10), or insurance (p=0.06). Patients in the control group were matched by both attending and location to patients in the sample group; all patients in this study were evaluated at one of three separate satellite locations within a single institution by 10 different retina specialists (**Table 1**).

### Anti-VEGF agent selection

In the 106 patients within the sample population, 22 (20.8%) patients received a sample injection of ranibizumab and 84 patients (79.2%) received a sample injection of aflibercept. At the second injection visit for the sample population (n=106), 2 patients (1.9%) received bevacizumab, 21 patients (19.8%) received ranibizumab, and 83 patients (78.3%) received aflibercept. At the third injection visit (n=97), 3 patients (3.1%) received bevacizumab, 20 patients (20.6%) received ranibizumab, and 74 received (76.3%) received aflibercept. At the fourth injection visit (n=86), 4 patients (4.7%) received bevacizumab, 16 patients (18.6%) received ranibizumab, and 66 patients (76.7%) received aflibercept. At one year (n=61), 0 patients (0%) received bevacizumab, 14 patients (23.0%) received ranibizumab, and 47 (77.0%) received aflibercept (**Table 2**).

**Table 2:** Anti-VEGF agent selection at each injection visit and at one year.

|  |  |  |  |  |  |
| --- | --- | --- | --- | --- | --- |
|  | **Injection 1** | **Injection 2** | **Injection 3** | **Injection 4** | **One year** |
| **Samples<sup>a</sup>** |  |  |  |  |  |
| Number of patients | 106 | 106 | 97 | 86 | 61 |
| Bevacizumab | 0 (0%) | 2 (1.9%) | 3 (3.1%) | 4 (4.7%) | 0 (0%) |
| Ranibizumab | 22 (20.8%) | 21 (19.8%) | 20 (20.6%) | 16 (18.6%) | 14 (23.0%) |
| Aflibercept | 84 (79.2%) | 83 (78.3%) | 74 (76.3%) | 66 (76.7%) | 47 (77.0%) |
| Number of patients<br>adherent to first injection | NA | 102 (96.2%) | 93 (95.9%) | 79 (91.9%) | 57 (93.4%) |
| **Controls<sup>a</sup>** |  |  |  |  |  |
| Number of patients | 106 | 106 | 104 | 99 | 74 |
| Bevacizumab | 43 (40.6%) | 41 (38.7%) | 43 (41.3%) | 40 (40.4%) | 25 (33.8%) |
| Ranibizumab | 18 (17.0%) | 19 (17.9%) | 17 (16.3%) | 16 (16.2%) | 10 (13.5%) |
| Aflibercept | 45 (42.4%) | 46 (43.4%) | 44 (41.5%) | 43 (43.4%) | 39 (52.7%) |
| Number of patients<br>adherent to first injection | NA | 104 (98.1%) | 98 (94.2%) | 94 (94.9%) | 65 (87.8%) |
| **Chi-Square Test for Samples vs. Controls** |  |  |  |  |  |
| P value | **<0.001** | **<0.001** | **<0.001** | **<0.001** | **<0.001** |

<sup>a</sup> All percentages are calculated as a proportion of number of patients at that injection visit.

For the first injection in the control population (n=106), 43 patients (40.6%) initially received bevacizumab, 18 patients (17.0%) received ranibizumab, and 45 patients (42.4%) received aflibercept. At the second injection visit for the control population (n=106), 41 patients (38.7%) received bevacizumab, 19 patients (17.9%) received ranibizumab, and 46 patients (43.4%) received aflibercept. At the third injection visit (n=104), 43 patients (41.3%) received bevacizumab, 17 patients (16.3%) received ranibizumab, and 44 received (41.5%) received aflibercept. At the fourth injection visit (n=99), 40 patients (40.4%) received bevacizumab, 16 patients (16.2%) received ranibizumab, and 43 patients (43.4%) received aflibercept. At one year (n=74), 25 patients (33.8%) received bevacizumab, 10 patients (13.5%) received ranibizumab, and 39 patients (52.7%) received aflibercept (Table 2).

There were relatively high adherence rates to the first injection for a patient’s second, third, and fourth injections as well as at one year for both the patients who were initially started on a sample (96.2%, 95.9%, 91.9%, and 93.4% respectively) and for control patients (98.1%, 94.2%, 94.9%, 87.8%), as seen in **Table 2**.

There was a statistically significant difference in the distribution of anti-VEGF injections given to the sample group and to the control group at each injection visit (p<0.001 for each visit, **Table 2**). **Figure 1** demonstrates the percentage of each anti-VEGF agent at each injection visit. Bevacizumab usage was significantly lower by chi-square test in samples relative to controls in the first four injections (p<0.001 for each) and at 1 year (p<0.001). Ranibizumab usage was not significantly different in samples relative to controls across the first four injections (p=0.48, p=0.73, p=0.44, p=0.66) and at one year (p=0.15). Aflibercept use was significantly increased in samples relative to controls during the first four injections (p<0.001 for each) and at one year (p<0.001).

**Figure 1:**
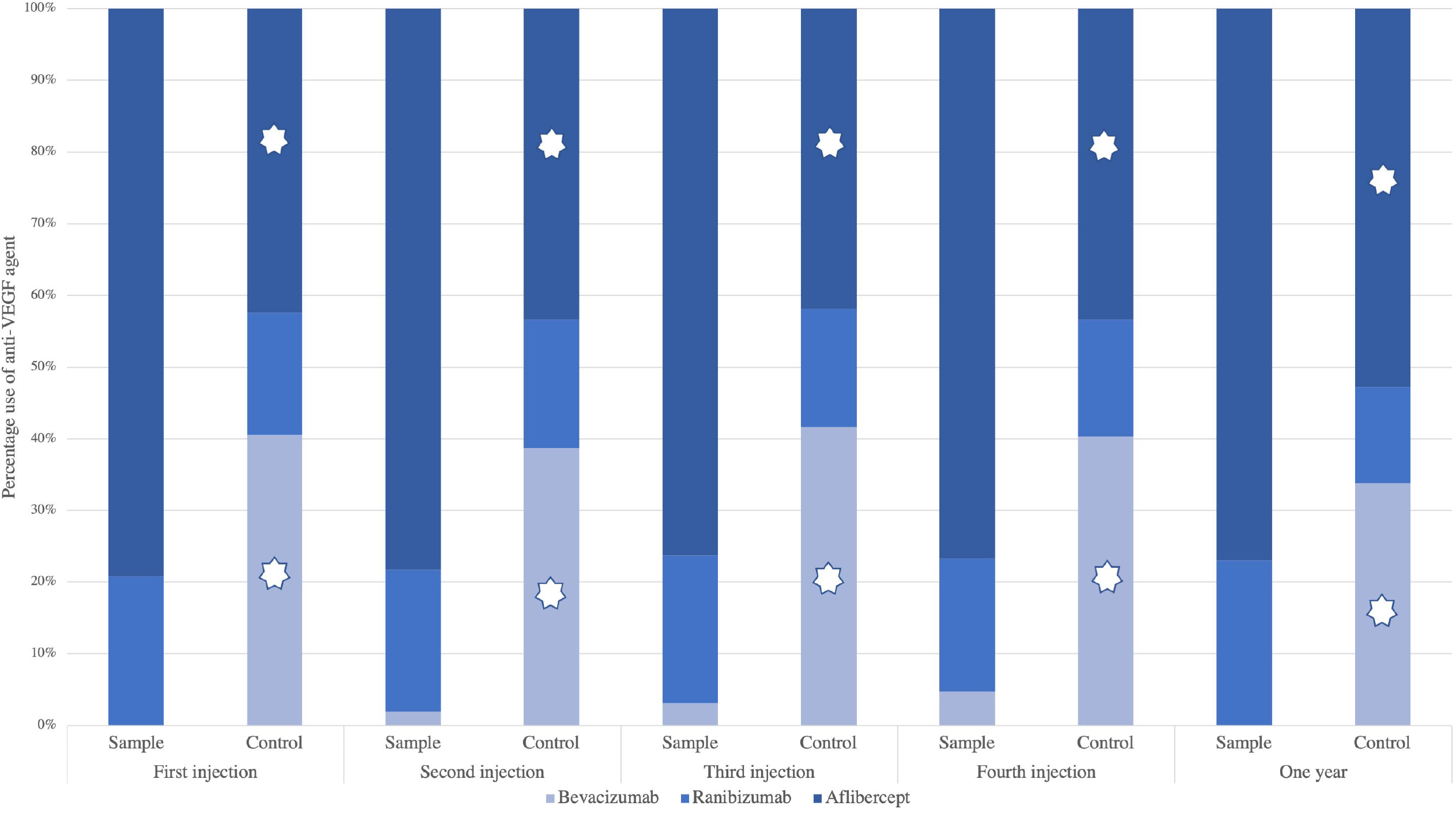
Percentage use of anti-VEGF agents at each visit for sample and control groups * denote p<0.001

### Time to injection from diagnosis and first retina visit

Time of the first visit with a retina specialist for evaluation of nvAMD to first anti-VEGF injection was significantly different between the sample and control groups (p=0.02), with a mean of 0.31 ± 1.84 days for the sample population and 1.19 ± 3.38 days for the control population (**Table 3**).

**Table 3:** Time to first injection for sample and control group.

| **Table 3: Time to first injection for sample and control group** |  |  |  |
| --- | --- | --- | --- |
|  | **Sample population**<br>**(n=106)** | **Control**<br>**population**<br>**(n=106)** | **P**<br>**value** |
| **Number of days from first visit with a**<br>**retinal specialist to first injection (mean,**<br>**SD)** | 0.31 ± 1.84 | 1.19 ± 3.38 | **0.02** |

### Visual acuity outcomes

Snellen visual acuity (VA) was recorded at each injection visit. Snellen VA was converted to logMAR form. There was no significant difference between logMAR acuity in the sample group and the control group during any of the injection visits. There was no significant difference in the improvement in logMAR acuity between the sample and control group from the first to the fourth visit (p=0.99, **Table 4**).

**Table 4:** Visual acuity of sample and control populations at each injection visit.

|  |  |  |  |
| --- | --- | --- | --- |
|  | **Sample population** | **Control population** | **P value** |
| **Visual Acuity (LogMAR) at first visit (mean, SD)** | 0.552 (0.369) | 0.504 (0.305) | 0.34 |
| **Visual Acuity (LogMAR) at second visit** | 0.440 (0.330) | 0.486 (0.299) | 0.40 |
| **Visual Acuity (LogMAR) at third visit (mean, SD)** | 0.443 (0.305) | 0.477 (0.321) | 0.47 |
| **Visual Acuity (LogMAR) at fourth visit (mean, SD)** | 0.441 (0.238) | 0.442 (0.224) | 0.99 |
| **Change in Visual (LogMAR) between first and fourth Visit (mean, SD)** | -0.106 (0.577) | -0.045 (0.250) | 0.48 |

### Bevacizumab use among attendings

Ten attendings contributed an equal number of sample and control patients to our study. In the sample population, rates of bevacizumab use were low (ranging from 0% to 4.0%), with 6/10 attendings never using bevacizumab after initial injection with a sample medication. In the control population, bevacizumab usage was higher, ranging from 15.4% to 100% among all injections examined in our study **(Table 5)**.

**Table 5:** Rate of bevacizumab usage among attendings within sample and control groups.

| Attending | Number of patient eyes contributed in each group per attending | Samples |  | Controls |  |
| --- | --- | --- | --- | --- | --- |
|  |  | Rate of bevacizumab usage for first injection | Rate of bevacizumab usage for total injections | Rate of bevacizumab usage for first injection | Rate of bevacizumab usage for total injections |
| 1 | 27 | 0/27 (0%) | 1/122 (0.8%) | 7/27 (25.9%) | 33/132 (25.0%) |
| 2 | 25 | 0/25 (0%) | 4/105 (3.8%) | 11/25 (44%) | 35/72 (48.6%) |
| 3 | 18 | 0/18 (0%) | 3/75 (4.0%) | 10/18 (55.6%) | 44/86 (51.1%) |
| 4 | 9 | 0/9 (0%) | 1/35 (2.9%) | 2/9 (22.2%) | 6/39 (15.4%) |
| 5 | 8 | 0/8 (0%) | 0/37 (0%) | 3/8 (37.5%) | 12/37 (32.4%) |
| 6 | 7 | 0/7 (0%) | 0/31 (0%) | 4/7 (57.1%) | 18/33 (54.5%) |
| 7 | 5 | 0/5 (0%) | 0/23 (0%) | 2/5 (40%) | 9/21 (42.9%) |
| 8 | 4 | 0/4 (0%) | 0/18 (0%) | 1/4 (25.0%) | 5/20 (25.0%) |
| 9 | 2 | 0/2 (0%) | 0/8 (0%) | 2/2 (100%) | 9/10 (90.0%) |
| 10 | 1 | 0/1 (0%) | 0/4 (0%) | 1/1 (100%) | 4/4 (100%) |

## Discussion

Our study found that in patients with nvAMD treated with anti-VEGF agents, there was a high adherence to the initial agent of choice for subsequent injections. Additionally, eyes of treatment naïve nvAMD patients who were initiated on a sample anti-VEGF agent had significantly lower rates of subsequent bevacizumab use compared to patients who did not receive an initial sample anti-VEGF agent despite the same attendings initiating treatment with bevacizumab among the control eyes.

We found that the use of sample medications for the initial treatment of nvAMD is associated with subsequent use of the associated brand name medication in the same eye. For physicians who initiated treatment with an anti-VEGF sample, there may be various reasons to maintain patients on that same agent for subsequent injections. There may be some physicians who always prefer brand-name agents in their personal practice patterns, and access to samples may not necessarily alter their behavior. However, the control population in our study was matched by both attending physician and location of treatment to the sample group. The physicians included in our study tended to use bevacizumab as an anti-VEGF agent more frequently in the control population group relative to the sample population. One possible reason might be a hesitancy in switching to an alternative agent if the initial medication initially showed efficacy for the patient. One might also imagine that switching to a cheaper, off-label medication for a patient’s second injection when they were started on an FDA-approved drug that is working may be difficult to explain to a patient. Regardless of the motivating factors, the inertia from using a sample initially leads to less use of lower cost bevacizumab in the majority of patients at least up to the one year follow-up period in this study.

There are several ethical and financial concerns with frequent medication sample usage, and some institutions have severely limited or banned the use of samples.^11^ One possible concern is that drug samples are a source of marketing that can have a major influence on physician prescribing behavior.^12^ Previous studies have found that sample availability can lead physicians to prescribe drugs that are different from their preferred drug choice in an attempt to reduce costs to the patient.^5^

Furthermore, samples may contribute to rising health care costs by increasing the subsequent use of more expensive products.^13,14^ The majority of anti-VEGF injections are for nvAMD.^9^ Medicare Part B alone spends more than 3.5 billion dollars annually on anti-VEGF medications, with the amount steadily increasing.^8^ The decreased utilization of bevacizumab among eyes receiving samples has significant economic ramifications for the healthcare system, given the major cost differential between the off-label medication bevacizumab versus the brand name medications aflibercept and ranibizumab.

Samples can, however, provide benefits to both physicians and patients. Sample medications facilitate rapid, no-cost treatment for patients and allow physicians to initiate therapy in a timely manner without the concern of incurring the financial liability of insurance denials. With sample medications, patients are able to start treatment immediately while awaiting insurance authorizations. Samples also allow treatment for off-label conditions for which it can be difficult to obtain insurance approval. This is especially important in anti-VEGF medication use, as on-label medications account for significant financial cost to physicians, clinics, and potentially patients if uncovered.

Our study found a small, though statistically significant, difference in the mean number of days from a nvAMD patient’s first visit with a retina specialist and their first injection for patients who received an initial sample compared to those who did not. Many of the major clinical trials (such as MARINA, ANCHOR, and VIEW1 and 2) demonstrating the efficacy of anti-VEGF use in nvAMD included only patients with recent or newly active exudation.^15^ Prompt treatment with anti-VEGF agents in the setting of recent conversion is important to maximize clinical and VA outcomes, and thus sample use is truly beneficial to patients as time sensitive treatment can be initiated without concern for insurance denials of a high cost drug.

One potential solution that may reduce healthcare spending on anti-VEGF medications while preserving urgent access to vision-saving medication and physician choice/patient autonomy would be for insurance companies to waive PAs for bevacizumab while samples are still available for physician use in clinics. In the American Medical Association physician survey in 2020, 94% of physicians reported care delays secondary to PA requirements.^16^ Waiving PAs may allow patients to have rapid access to a medication while also decreasing the administrative burdens on physicians and clinics. As things stand now, Repka et al. recently reported that bevacizumab use has declined over the past 7 years across all included insurance payors according to data from the Intelligent Research in Sight (IRIS) registry despite an increase in tools such as prior authorization and step therapy.^17^

Optimal anti-VEGF agent choice for nvAMD continues to be debated at length, and it is well beyond the scope and goal of our study. We are fortunate that each treatment is an effective one for nvAMD and recognize the importance of allowing both physician and patient autonomy in treatment choice. There are many clinical situations when a specific medication may not be the ideal anti-VEGF agent of choice, including limited efficacy in some patients, medication allergies, patients who are unwilling to consent to use of an off-label drug therapy, cost to patients, or physician/patient preference. Furthermore, we emphasize that the goal of our study is not to study the efficacy of a particular anti-VEGF agent, nor is it to advocate for on-label or off-label medications, but to understand the potential association of anti-VEGF sample medication use with ultimate anti-VEGF agent selection. Lastly, we do not advocate for a ban on anti-VEGF medication samples, as their availability has some benefits for patients. In all these situations, the availability of multiple anti-VEGF agents and sample medications to test their efficacy is valuable to both patient and clinician. Minimizing barriers to similarly efficacious, but lower cost options such as bevacizumab could relieve administrative burdens on physicians and staff, lead to lower healthcare spending, and may ultimately help patients obtain prompt treatment.

Limitations to this study include its retrospective nature. Additionally, the patient population and retina specialists are from a single institution and may not be representative of nationwide practice patterns. Another limitation is that sample utilization was only examined in patients who received an anti-VEGF sample as their initial treatment. Patients may have received samples is in the middle of their injection course (for example, when a physician is considering whether to switch agents for greater efficacy), and these patients were not included in our study.

Our study demonstrates that when a sample anti-VEGF is the first treatment, there is persistent use of the more expensive medication in subsequent injections at least up to one year. Ophthalmologists should be cognizant of how samples may impact future medication use whether directly or indirectly due to administrative, clinical, and patient factors. Payors should also consider how requiring prior authorizations may paradoxically increase health care costs, which is detrimental to the payors directly as well as society as a whole.

## Data Availability

Data are available upon reasonable request.

## A. Funding/support

None

## B. Financial Disclosures

1. JBM is a consultant for Alcon, Allergan, Carl Zeiss, Sunovion, and Genentech.
2. DH is a consultant for Allergan, Genentech, Omeicos Ophthalmics, and has received financial support from Lions VisionGift, Commonwealth Grant, and Macular Society.
3. LAK has received financial support from National Eye Institute and CureVac AG, has financial arrangement with Pykus Therapeutics, and holds patents through Massachusetts Eye and Ear.
4. DGV is a consultant for Valitor, Olix Pharmaceuticals, and has received financial support from National Eye Institute, Research to Prevent Blindness, Loefflers Family Foundation, Yeatts Family Foundation, and Alcon Research Institute.
5. DE is a consultant for Alcon, Allergan, Dutch Ophthalmics, Genentech, and Glaukos. He has a financial relationship with Alderya Therapeutics and Pykus Therapeutics, and has received financial support from Neurotech Pharmaceuticals.
6. ESG has a financial relationship with Auro Biosciences, Astellas Pharma, and Valeant Pharmaceuticals.
7. JWM is a consultant for Genentech/Roche, KalVista Pharmaceuticals, Ltd., Sunovion, Heidelberg Engineering, LifeBiosciences, Inc; has received financial support from Lowy Medical Research Institute, Ltd.; has royalties with Valeant Pharmaceuticals/Mass Eye and Ear, ONL Therapeutics, LLC; Aptinyx, Inc; and holds a patent through Valeant Pharmaceuticals/Mass Eye and Ear, ONL Therapeutics, LLC.
8. DMW holds a patent through Massachusetts Eye and Ear.
9. RP was a consultant for Anthem Blue Cross and Blue Shield.
10. All other authors (KMW, TB, SP, EC, JK, MEA, LHH, RH, SM, NAP, LS) have reported that they have no relationships relevant to the contents of this paper to disclose.

## C. Other acknowledgments

The authors would like to thank Stephanie Gooltz and Fran McDonald for their help on this project.

## Notes

### Funding Statement

There is no funding support to report.

### Author Declarations

This study was approved by the institutional review board of the Massachusetts General Hospital and Partners Healthcare (IRB # 2021P000301) and is compliant with the Declaration of Helsinki and Health Insurance Portability and Accountability Act regulations.

